# The Resynchronization Effect in Left Bundle Branch Pacing can be Evaluated Non-invasively with the Implementation of Lead V8

**DOI:** 10.64898/2025.12.03.25341597

**Authors:** Vadivelu Ramalingam, Johan van Koll, Peter Tai, Arno Fennema, Vidal Essebag, Atul Verma, Uyên C. Nguyên, Joost Lumens, Justin G.L.M. Luermans, Kevin Vernooy, Jacqueline Joza

## Abstract

**Background:** Left bundle branch area pacing (LBBAP) is increasingly being used for cardiac resynchronization therapy (CRT); however an additional left ventricular (LV) lead is required when resynchronization remains incomplete. This study evaluates whether lead V8 can provide a simple, non-invasive marker of persistent posterolateral LV delay during LBBAP.

**Methods:** Consecutive patients undergoing LOT-CRT implantation were included. Standard 12-lead ECGs were obtained with the V5 electrode repositioned to the V8 location. Local activation time was measured as the steepest negative downslope of the QRS (negative derivative activation time, NDAT) and compared with the LV electrical delay (LVED) determined from the LV lead during LBBAP.

**Results:** Thirty consecutive patients undergoing LOT-CRT implantation were included with a total of 106 ECG recordings with corresponding LVED measurements. The NDAT-V8 demonstrated a strong correlation with the LVED during intrinsic conduction (r = 0.95) and during all LBB pacing subtypes: combined r = 0.922; mean difference 2.5 ± 8 ms; RV septal pacing, r = 0.89; LV septal pacing, r = 0.92; non-selective LBBP, r = 0.91; and selective LBBP, r = 0.81. The correlation of LVED during intrinsic conduction and all LBBAP pacing subtypes was significantly weaker for NDAT-V6 and the RWPT in V6 and V8 (r=0.681, 0.626 and 0.726, respectively).

**Conclusion:** The NDAT-V8 provides a reliable non-invasive surrogate for the LV posterolateral wall delay during LBBAP, outperforming NDAT-V6 and the RWPT in V6 and V8 and establishes the groundwork for future studies evaluating NDAT-V8 as a tool to guide adequate resynchronization during LBBAP.

**Clinical Perspective:** *What is Known:*

- Although left bundle branch area pacing (LBBAP) is increasingly being used as an alternative resynchronization strategy, complete resynchronization is not always achieved.
- The negative derivative activation time in lead V8 (NDAT-V8) has been shown to provide a non-invasive marker of the left ventricular electrical delay– also referred to as QLV - at the LV postero-lateral wall in patients with left bundle branch block, intraventricular conduction delay, and right bundle branch block.

*What the Study Adds:*

- This study shows that the NDAT-V8 provides a non-invasive surrogate for the left ventricular posterolateral wall delay during LBBAP.
- This study establishes the groundwork for future studies evaluating NDAT-V8 as a tool to guide adequate resynchronization therapy during LBBAP to determine when the addition of a coronary sinus lead is needed.

## Introduction

Cardiac resynchronization therapy (CRT) is a well-established therapy for patients with heart failure (HF) with reduced ejection fraction and prolonged QRS duration, having shown improvements in symptoms and reductions HF hospitalizations and mortality.(1) Left bundle branch area pacing (LBBAP) is increasingly being used as an alternative resynchronization strategy, aiming to directly restore physiologic ventricular activation by engaging the native conduction system.(2) Observational and the early small randomized studies are promising and have demonstrated favorable outcomes with LBBAP for CRT. (3–6)

Complete resynchronization is, however, not always achievable with LBBAP.(7) Attaining only LV septal pacing, rather than true conduction system capture, has been associated with suboptimal outcomes.(8) Moreover, achieving only deep septal capture is particularly suboptimal in patients requiring resynchronization therapy.(9) These limitations have fueled a growing emphasis on the personalization of resynchronization strategies, tailoring therapy to the underlying conduction substrate.(10) For example, patients with left bundle branch block (LBBB) that can be corrected by LBBP may derive greater benefit from LBBP-CRT, whereas those with predominant IVCD may be better suited to BiV-CRT. However, in practice the decision is rarely so straightforward. Uncertainty often arises regarding which therapy to initiate, or whether a left bundle branch pacing optimized – CRT (LOT-CRT) strategy, which includes implantation of an additional LV lead, may offer the greatest benefit.

Regardless of the chosen approach, achieving complete resynchronization remains the ultimate therapeutic goal. Recently, the negative derivative activation time in lead V8 (NDAT-V8) has been shown to provide a non-invasive marker of the left ventricular (LV) electrical delay (LVED) – also referred to as QLV - at the LV postero-lateral wall in patients with LBBB, intraventricular conduction delay (IVCD), and right bundle branch block (RBBB). It was hypothesized that the LV posterolateral wall delay in LBBAP could serve as an intra-procedural marker to guide the resynchronization effect by LBBAP. Therefore, the aim of this study was to determine whether NDAT-V8 may be used as a non-invasive marker of the LV postero-lateral wall electrical delay to guide effective resynchronization during LBBAP and identify when the addition of an LV lead may be warranted.

## Methods

### Study Design and Population

Consecutive patients undergoing LOT-CRT implantation underwent V8 lead recording at both the McGill University Health Centre (MUHC, Montreal, Canada) and the Maastricht University Medical Centre (MUMC, Maastricht, the Netherlands) between March and November 2025. Patients in whom the LV lead was not positioned in a posterolateral or lateral vein were excluded. The indication for LOT-CRT was determined at the discretion of the implanting physician, and procedures were performed according to clinical practice. The study protocol was approved by corresponding institutional ethical review boards.

### ECG lead V8 positioning

Standard surface ECG recordings were recorded during the procedure using electrophysiology (EP) recording systems (CardioLab 6.9.6. GE Medical in Montreal, and Boston Scientific LabSystem PRO EP 4.2.0 in Maastricht) which enabled 12-lead ECG-, as well as filtered- (30Hz – 250Hz) and unfiltered EGM-recordings obtained from the electrodes on the LV lead as well as the LBBP lead. Limb and precordial leads were positioned according to standard practice, except for lead V5. This lead was positioned at the V8 position, on the back, ±3 centimeters below the tip of the left scapula, in the same horizonal plane as lead V6 (figure 1).(11) Practically, this corresponds to the midscapular line, directly posterior to leads V4-V6. At the final implantation sites, surface ECGs and unipolar EGMs were recorded during intrinsic rhythm and LBBAP.

**Figure 1.**
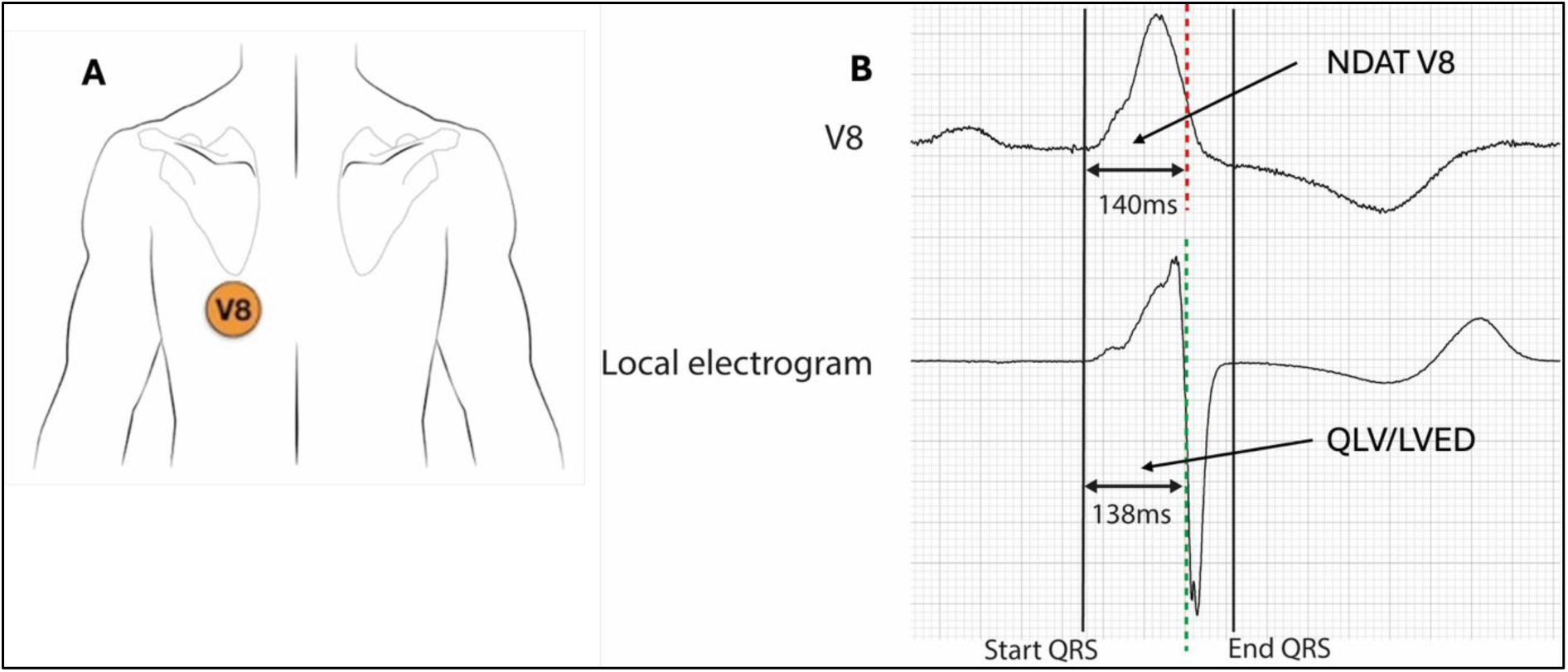
A. Lead V5 has been moved to the V8 position on the back, at the tip of the left scapula, in the same horizonal plane as lead V6. B. The NDAT and LVED interval measurement from onset of QRS to steepest negative downslope on the unipolar electrogram. (NDAT-V8 (top) red vertical dashed line, and LVED (bottom) green vertical dashed line). Abbreviations. NDAT: negative derivative activation time. LVED: left ventricular electrical delay.

### ECG, Signal Analysis and NDAT measurements

The time of maximum negative derivative (ND) of the unipolar EGMs was used to determine the moment of local electrical tissue activation.(12, 13) The LV electrical delay (LVED) was defined as the interval between the earliest QRS onset (during intrinsic rhythm) or pacing stimulus (during paced rhythm) in any surface lead and the above described EGM-based moment of local activation, recorded from each LV lead electrode. In this study, *LV electrical delay (LVED)* is used in place of *QLV*, as the latter does not accurately describe measurements obtained during ventricular pacing. For each patient, the LVED interval was defined as the longest LVED interval across all available LV lead electrodes.

As the precordial leads represent unipolar recordings, the negative derivative activation time (NDAT) of the QRS complex was obtained from lead V1, V2, V3, V4, V6 and V8 as the interval between earliest QRS onset in any surface lead and the NDAT of each lead. The time difference between the LVED and NDAT during intrinsic rhythm, as well as the LVED and NDAT during LBBAP were determined for leads V6 and V8. The R wave peak times in V6 and V8 were also calculated during LBBAP. An LVED interval longer than the NDAT yields a positive value, whereas a LVED interval shorter than the NDAT yields a negative value. All LVED and NDAT measurements were performed manually on the EP recording systems at a paper speed of 50 to 200mm/s.

### LOT CRT procedure

The LV, LBBAP, and right atrial leads were implanted according to clinical practice.(2, 14–17) The targeted LV lead position was the LV posterolateral wall with the longest LVED. The LV lead was placed in a LV posterolateral mid-basal region in all patients. LBBAP implantation followed a stepwise strategy, beginning at the proximal left bundle and progressing distally if capture was not achieved. Successful LBB capture was defined according to established criteria, with demonstration of transition serving as the primary indicator of capture.(18) Additional activation criteria were used as supportive evidence. In the presence of septal scar or slow conduction, functional deep septal capture (in the presence of transition) or LVSP was accepted, following repeated attempts at positioning.

Surface ECGs and unipolar EGMs were recorded during intrinsic rhythm, and across all available LBBAP types: deep septal pacing (DSP), LV septal pacing (LVSP), and at the final lead position with high output (non-selective LBBP, nsLBBP) and low output (LVSP or selective LBBP, sLBBP), RV septal pacing (pacing from the ring of the LBBAP lead). Pacing was performed in unipolar fashion. The LVED was measured from the LV lead during intrinsic rhythm and across all available LBBAP types. The latest LVED was chosen for the final analysis. Understandably, it was not possible to achieve all LBBAP configurations in every patient.

### Statistical methods

Continuous variables are reported as mean ± standard deviation (SD) or median with interquartile range [IQR]. Categorical variables are presented as counts (n) and percentages (%). Differences between the NDAT V8 and the latest LVED across various pacing types were analysed using mean or median of all individual differences. The relationship between the NDAT-V8 and the latest LVED was assessed using linear regression analysis, with the calculation of Pearson’s correlation coefficient (*r*). Agreement between LVED and NDAT measurements was evaluated using Bland–Altman analysis and Lin’s concordance correlation coefficient (CCC) to assess systematic bias and variation across the measurement range. A uniform distribution within the 95% limits of agreement was interpreted as evidence of consistent agreement and absence of proportional bias. The CCC was employed as it accounts for both correlation and accuracy relative to the line of identity (y = x). A two-sided p < 0.05 was considered statistically significant. Analyses were performed using Python 3.12.12.

## Results

### Patient Characteristics

A total of 30 consecutive patients underwent LOT-CRT implantation and were included in the analysis. The majority of the implants (n=27/30, 90%) were *de novo* procedures, while 3 (10%) were upgrades. The baseline characteristics are summarized in table 1. The intrinsic QRS morphology was Strauss-criteria defined LBBB in 17 (57%), intraventricular conduction delay (IVCD) in 3 (10%), right bundle branch block (RBBB) in 7 (23%), narrow QRS in 2 (7%), and paced rhythm in 1 (3%). The mean age of the study population was 76±9 years and 57% were male. The baseline LVEF was 34±12% and 15 (50%) patients had a diagnosis of ischemic cardiomyopathy. Sinus rhythm was present in 17 patients (57%), atrial fibrillation (AF) in 12 (40%) and 1 patient was ventricular pacing-dependent (3%).

**Table 1.**
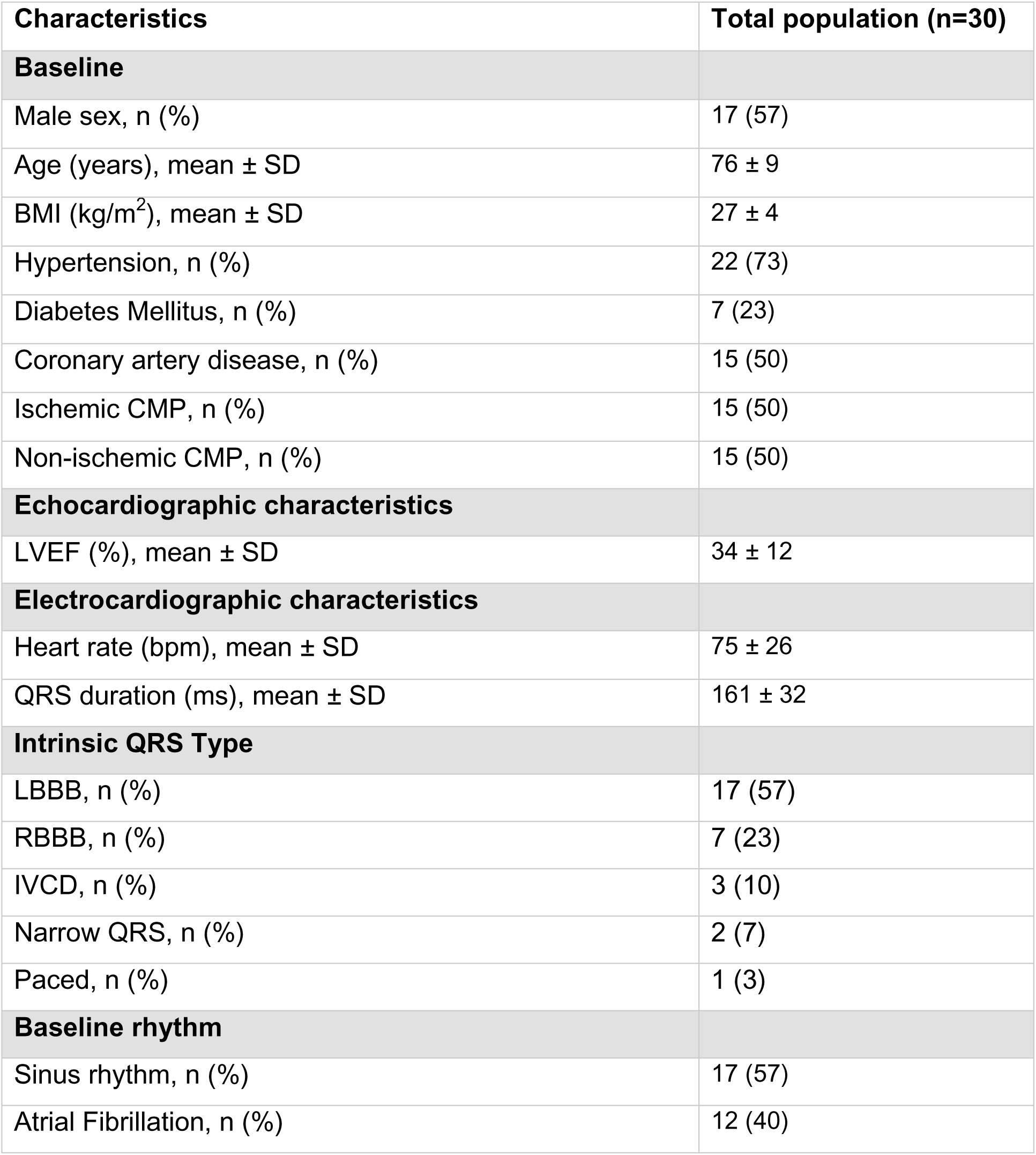
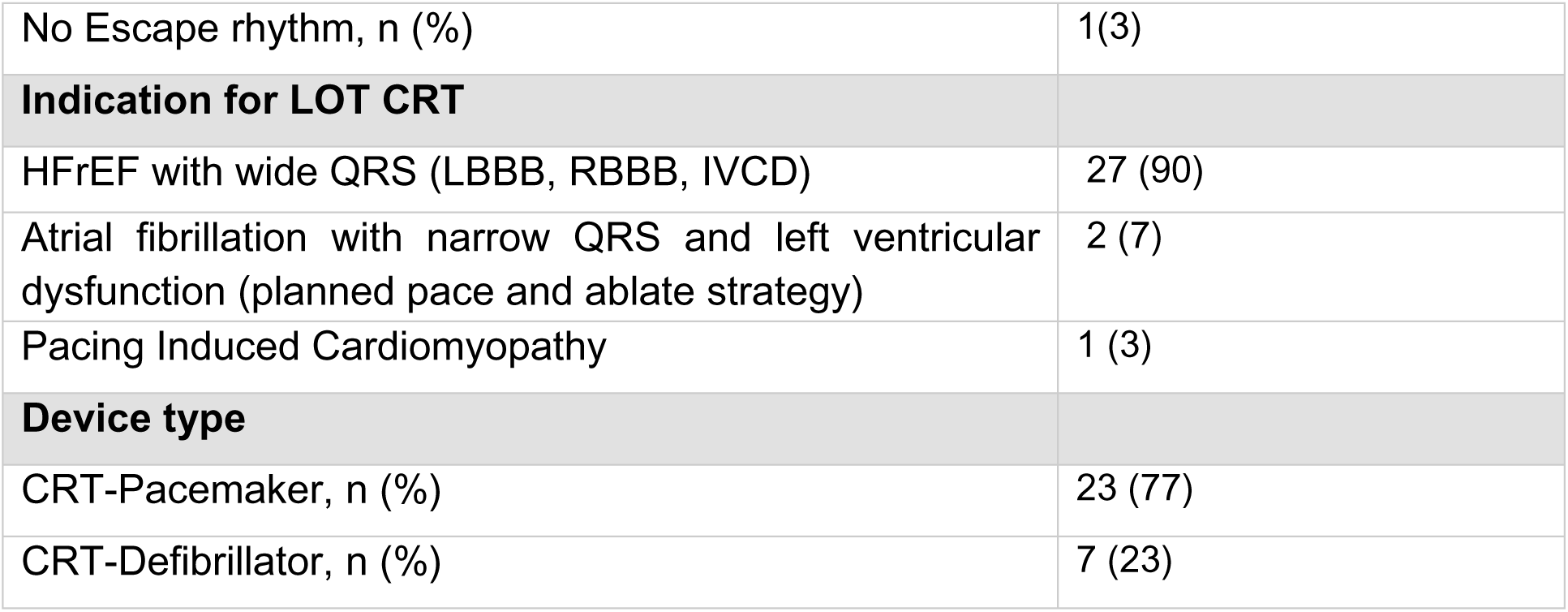
Baseline patient characteristics. AF: Atrial Fibrillation; BMI: Body Mass Index; CMP: Cardiomyopathy; CRT: Cardiac Resynchronization Therapy; HFrEF: Heart Failure with Reduced Ejection Fraction; IVCD: Intraventricular Conduction Delay; LBBB: Left Bundle Branch Block; LOT-CRT: Left Bundle Branch Optimized Cardiac Resynchronization Therapy; LVEF: Left Ventricular Ejection Fraction; PICM: Pacing Induced Cardiomyopathy; RBBB: Right Bundle Branch Block

### LBBP Implantation

Transition was observed in 21 (70%) where non-Selective to selective LBBP transition was observed in 18 (60%) and non-Selective LBBP to LVSP transition in 3 (10%). Where no transition was noted and no further evidence of left bundle branch capture was achieved, the result was LVSP in 6 (20%) and DSP in 3 (10%). The location of the lead was proximal in 10 (30%), posterior fascicular in 9 (30%), and anterior fascicular in 1 (3%) as per operator discretion. LBBP characteristics are summarized in table 2.

**Table 2.**
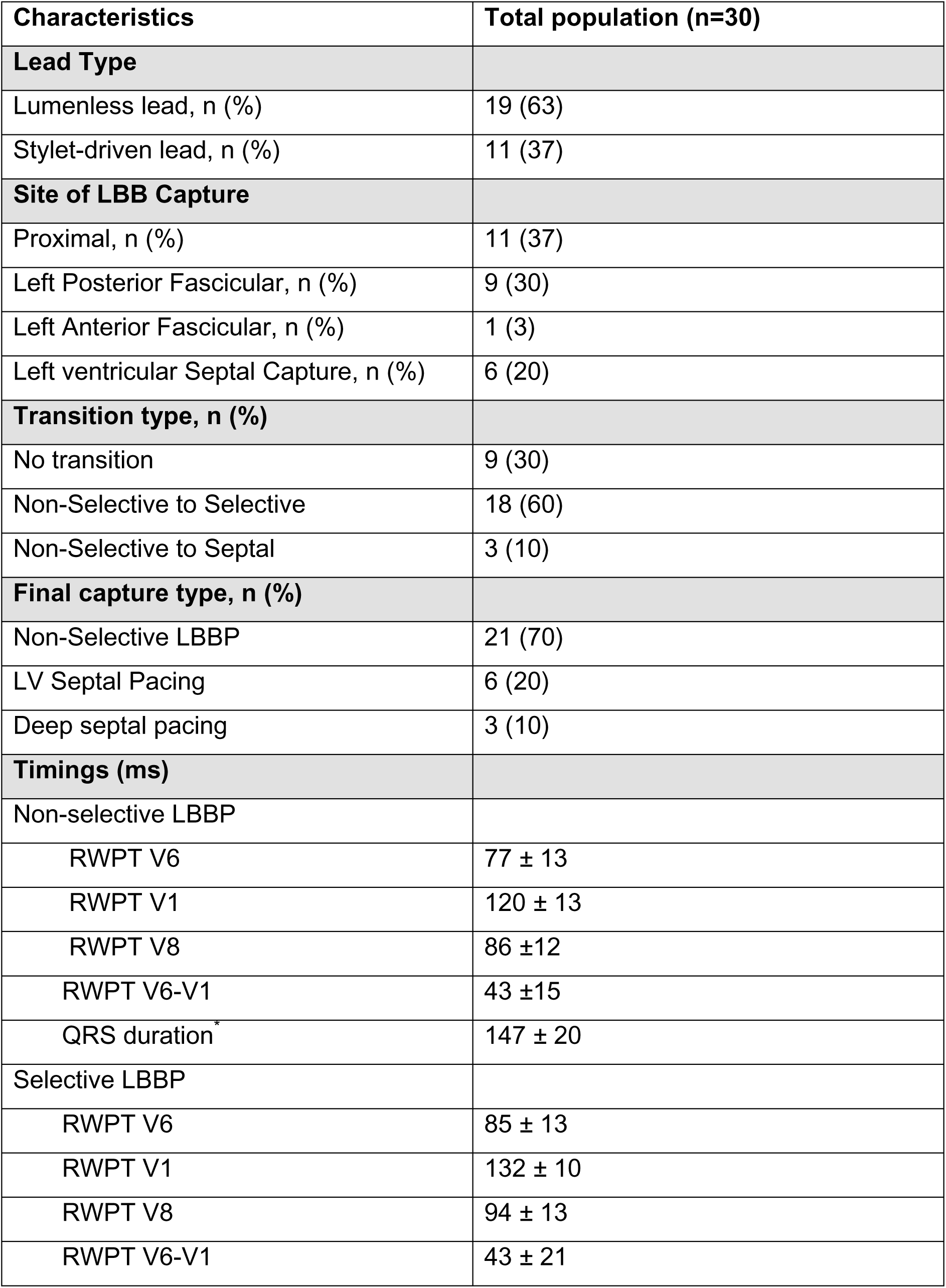
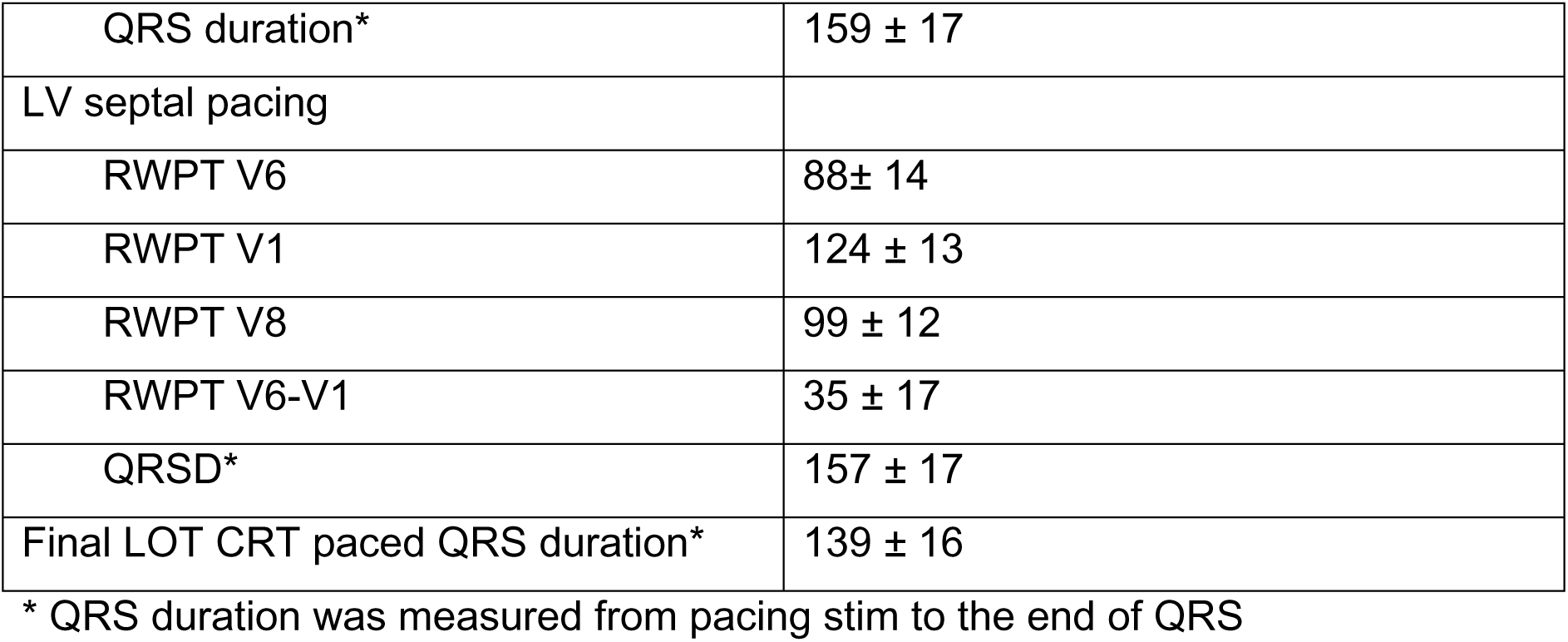
Characteristics of the left bundle branch pacing implant. Abbreviations: LBBB: left bundle branch; LV: left ventricular; RWPT: R-wave peak time

### NDAT and LVED measurements during LBBP pacing and intrinsic rhythm

A total of 106 ECG recordings with corresponding LVED measurements were obtained, some of which were obtained after achieving capture, and some during continuous pacing with continuous LVED recording. As a result, measurements included 29 intrinsic rhythm measurements, 22 RVSP, 8 DSP, 12 LVSP, 20 nsLBBP, and 15 sLBBP. There was close alignment between NDAT-V8 and LVED for intrinsic rhythm and across all LBBP subtypes. For the combined cohort of LBB pacing subtypes, the mean difference was 2.5±8ms, 1 ± 10 ms for intrinsic rhythm, 2 ± 8 ms for RVSP, -0.4 ± 8 ms for LVSP, 4.5 ± 8 ms for nsLBBP, and 4 ± 8 ms for sLBBP. In contrast, the LVED–NDAT V6 difference showed much larger offsets (combined:19 ±17 ms, 22 ± 20 ms, 14 ± 11 ms, 11 ± 9 ms, 23 ± 21 ms, and 20 ± 13 ms, respectively).

### V1-V8 Activation Time Plots

Activation time plots were constructed for the negative derivative of V1-V8, and to characterize the relationship between NDAT-V8 and the latest LVED during intrinsic rhythm and across LBBP subtypes. A typical activation time plot of one such patient with underlying LBBB who underwent LOT-CRT is depicted in figure 2. It clearly shows that the NDAT-V1 to V8 activation pattern during intrinsic LBBB mirrored that of RVSP. With septal penetration of the lead during DSP, the NDAT-V8 and LVED slightly shortened as compared to intrinsic and RVSP. With LVSP the NDAT-V8 further shortened but delayed activation of the RV developed as evidenced by a abrupt prolongation of the NDAT in V1. As the lead reached the LV subendocardium demonstration of nsLBBP and sLBBP was observed. The nsLBBP demonstrated further shortening in the NDAT-V8 and LVED times. The NDAT-V1 further prolonged with sLBBP by 15ms. There was near-perfect correlation between NDAT-V8 and LVED across all LBBP subtypes. The mean NDAT as well as LVED values across all LBBP subtypes of the entire cohort are noted in table 3.

**Figure 2.**
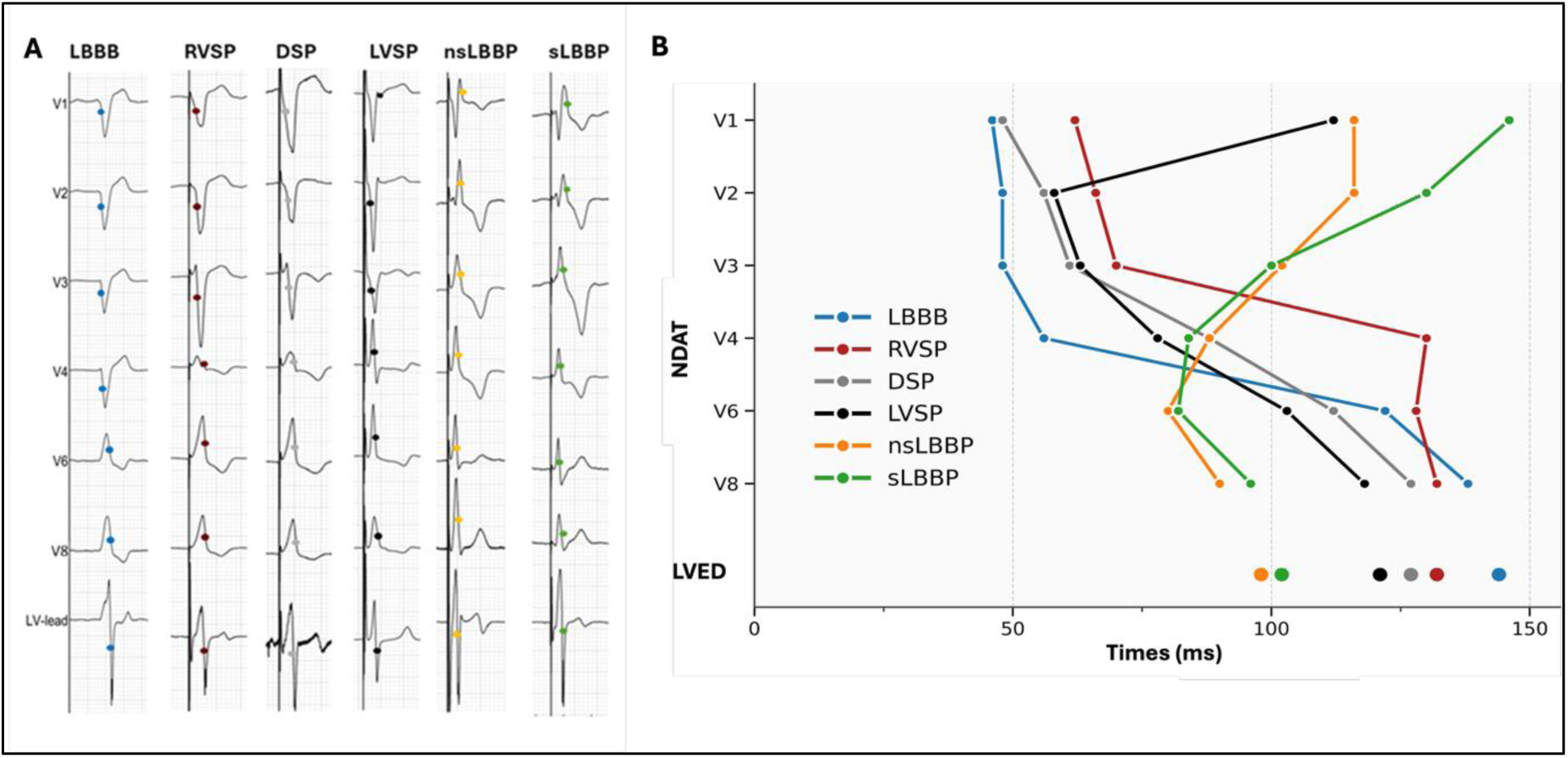
A. Activation time plot of a representative patient with baseline LBBB with the negative derivative activation time recorded for V1 to V8 across LBBP subtypes with corresponding LVED. B. Baseline LBBB seen in blue, RVSP in maroon, DSP in grey, LVSP in black nsLBBP in orange and sLBBP in green). The LVED times are represented by corresponding colored dots. Abbreviations: DSP: Deep septal pacing; LBBB: left bundle branch block; LBBP: left bundle branch pacing; LVED: left ventricular electrical delay; LVSP: Left ventricular septal pacing; NDAT: Negative derivative activation time; nsLBBP: non-selective left bundle branch pacing; RVSP: right ventricular septal pacing; sLBBP: selective left bundle branch pacing.

**Table 3.**
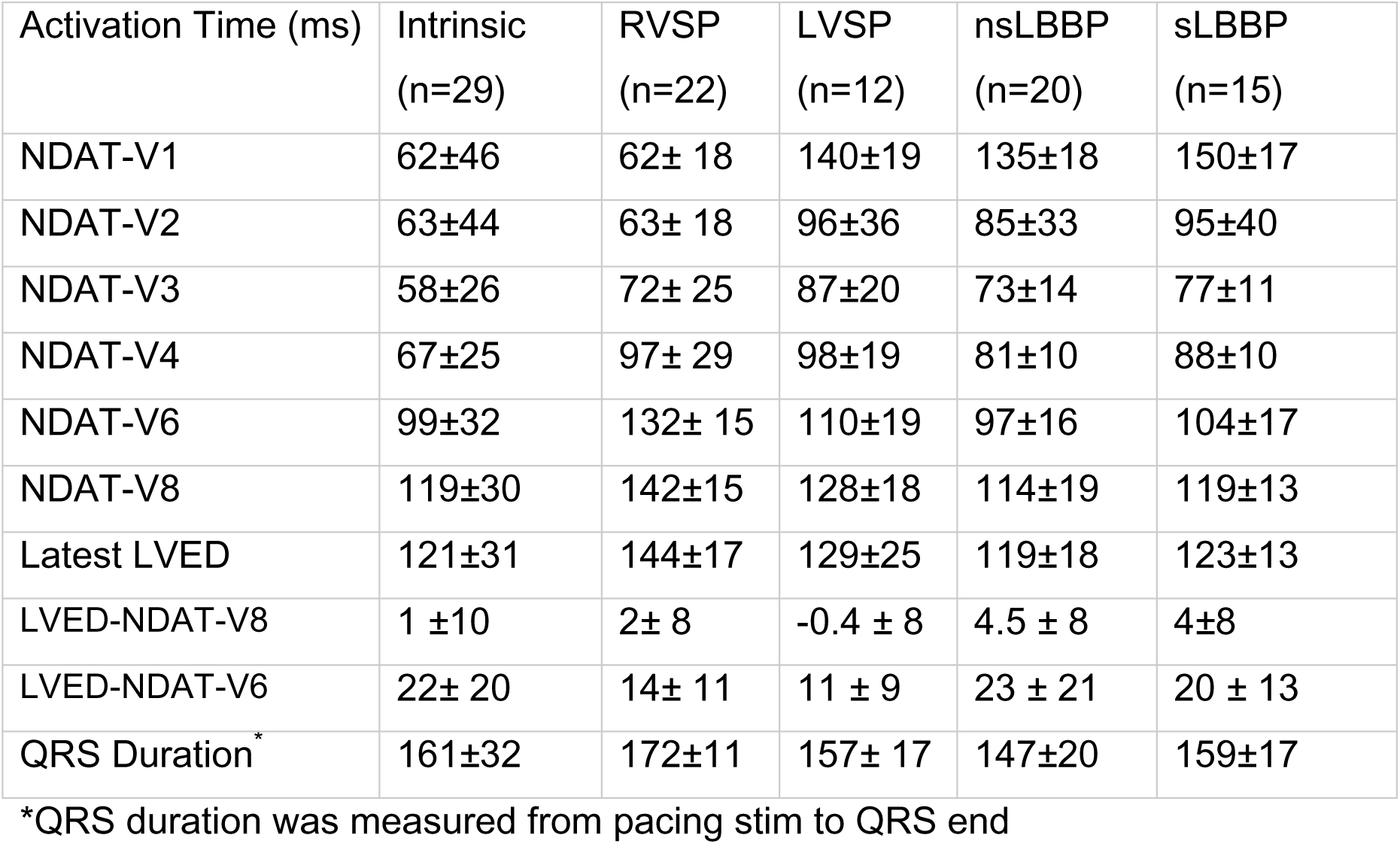
NDAT and LVED measurements across LBBP subtypes. Abbreviations: NDAT: negative derivative activation time; LBBP: left bundle branch pacing; LVED: Left ventricular electrical delay; LVSP: left ventricular septal pacing; nsLBBP: nonselective left bundle branch pacing; RVSP: right ventricular septal pacing; sLBBP: selective left bundle branch pacing

| Activation Time (ms) | Intrinsic<br>(n=29) | RVSP<br>(n=22) | LVSP<br>(n=12) | nsLBBP<br>(n=20) | sLBBP<br>(n=15) |
| --- | --- | --- | --- | --- | --- |
| NDAT-V1 | 62±46 | 62± 18 | 140±19 | 135±18 | 150±17 |
| NDAT-V2 | 63±44 | 63± 18 | 96±36 | 85±33 | 95±40 |
| NDAT-V3 | 58±26 | 72± 25 | 87±20 | 73±14 | 77±11 |
| NDAT-V4 | 67±25 | 97± 29 | 98±19 | 81±10 | 88±10 |
| NDAT-V6 | 99±32 | 132± 15 | 110±19 | 97±16 | 104±17 |
| NDAT-V8 | 119±30 | 142±15 | 128±18 | 114±19 | 119±13 |
| Latest LVED | 121±31 | 144±17 | 129±25 | 119±18 | 123±13 |
| LVED-NDAT-V8 | 1 ±10 | 2± 8 | -0.4 ± 8 | 4.5 ± 8 | 4±8 |
| LVED-NDAT-V6 | 22± 20 | 14± 11 | 11 ± 9 | 23 ± 21 | 20 ± 13 |
| QRS Duration* | 161±32 | 172±11 | 157± 17 | 147±20 | 159±17 |

\*QRS duration was measured from pacing stim to QRS end

### Correlation of NDAT-V8 with LVED

The NDAT-V8 correlated well with LVED across all LBBP subtypes (Pearson’s r=0.92, 95% CI 0.88–0.95; p<0.001) (figure 3A). In the pooled Bland–Altman analysis (figure 3B), LVED and NDAT-V8 demonstrated good overall agreement with a mean bias of +2.5±7.8 ms, yielding 95% limits of agreement from −12.8 to 17.8ms. There was no evidence of proportional bias (slope 0.048, p = 0.31), indicating consistent agreement between LVED and NDAT V8.

**Figure 3.**
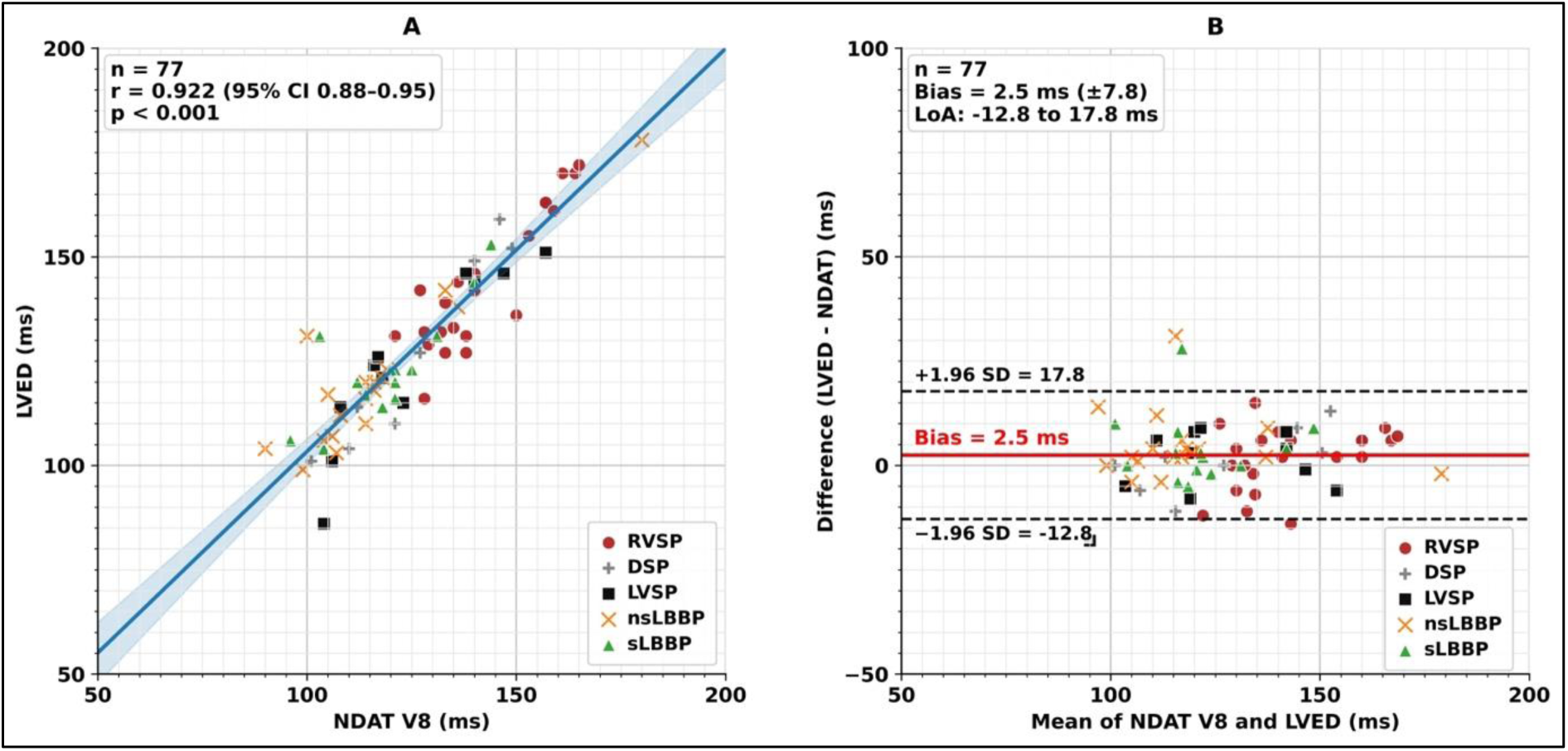
Correlation and Bland–Altman analyses of NDAT-V8 and LVED across pacing subtypes. 3A. Correlation scatter plot of NDAT-V8 (x-axis) vs LVED (y-axis), with the overall fit (solid blue line) with corresponding 95% CI (shaded light blue). Pearson’s r with 95% CI and p-value in the upper-left corner. Points are colour-coded by pacing subtype(RVSP, DSP, LVSP, nsLBBP, and sLBBP). 3B. Bland-Altman analysis of LVED–NDAT V8 . The mean bias of 2.5ms is represented by the red solid line and limits of agreement (LOA) ±1.96 SD are noted by the black-dashed lines. Abbreviations: DSP, deep septal pacing; LVED, left ventricular electrical delay; LVSP, left ventricular septal pacing; RVSP, right ventricular septal pacing; nsLBBP, non-selective left bundle branch pacing; sLBBP, selective left bundle branch pacing.

### Correlation and Concordance during individual LBBAP pacing modes

NDAT-V8 showed good correlation and concordance with LVED across all different pacing subtypes (figure 4A-D). The correlation (r) was 0.89 for RVSP, 0.92 for LVSP, 0.91 for nsLBBP and 0.81 for sLBBP (p<0.001). NDAT-V8 demonstrated the smallest mean bias relative to LVED (2.5±8ms), compared with NDAT-V6 (19 ± 17ms), RWPT-V6 (38±16.0ms), and RWPT-V8 (30±14ms) (figure 5). Overall, NDAT-V8 exhibited the lowest systematic error (bias closest to zero) and the narrowest dispersion of differences.

**Figure 4.**
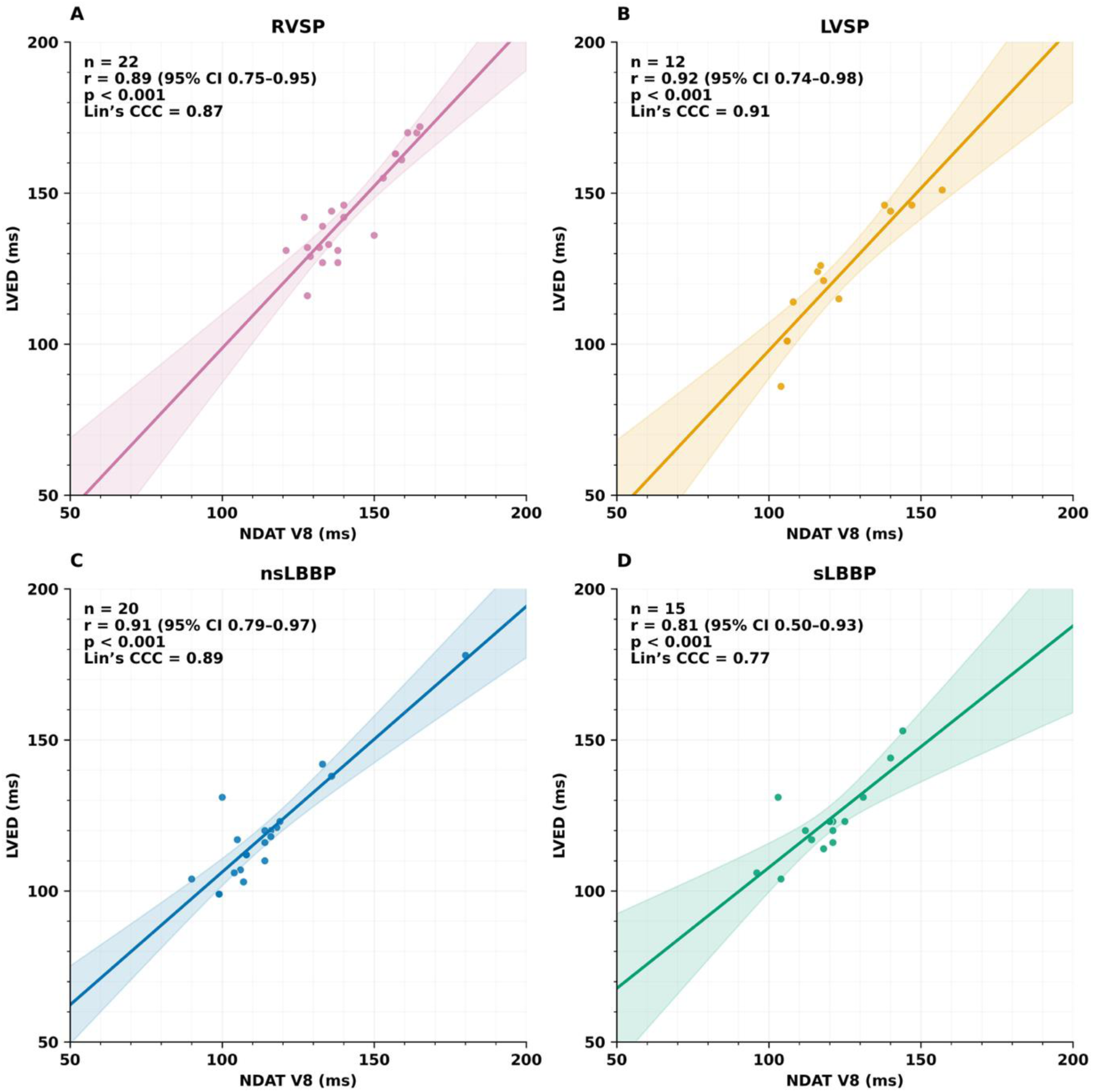
Correlation of NDAT-V8 with LVED across LBB pacing subtypes. (A) RVSP (B) LVSP (C) nsLBBP and (D) sLBBP. Each panel shows scatter points with a least-squares fit (solid line) and the 95% CI (shaded). Number of observations (n), Pearson’s r with its 95% CI and p-value, and Lin’s concordance correlation coefficient (Lin’s CCC).

**Figure 5.**
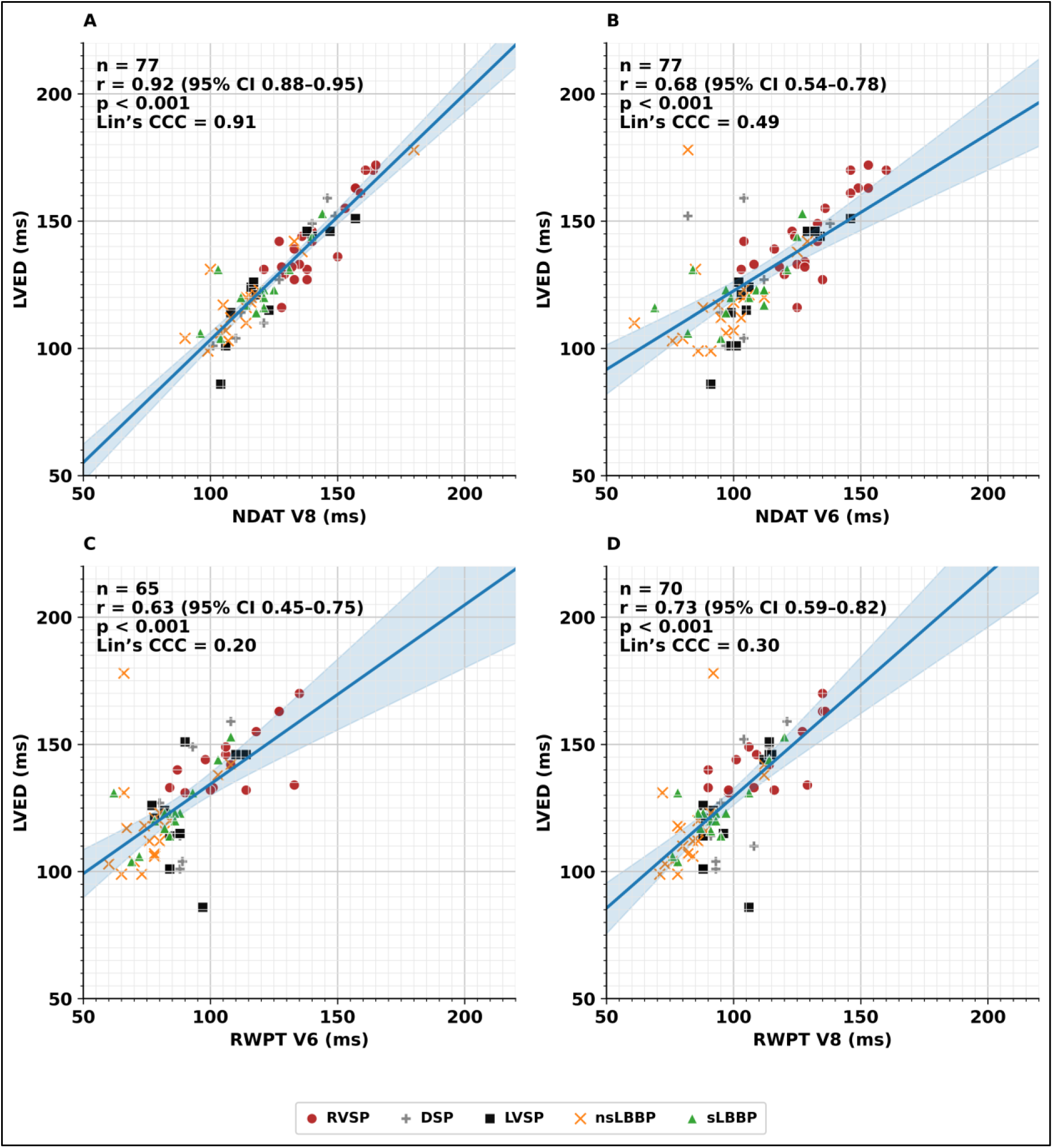
Correlation and concordance of activation times with the latest LVED across LBBP subtypes. 5A. NDAT-V8; 5B. NDAT-V6; 5C. RWPT-V6; 5D. RWPT-V8. The solid blue line is the least-squares regression; the translucent blue band denotes its 95% confidence interval. Points are symbol/color-coded by pacing mode (RVSP, DSP, LVSP, nsLBBP, and sLBBP). Lin CCC: Concordance Correlation Coefficient is also displayed.

## Discussion

This study demonstrates that the negative derivative activation time of the V8 electrode is a powerful noninvasive measurement to recognize LV posterolateral wall delay during LBBAP. The main findings are as follows: (1) There is a strong correlation between non-invasively measured NDAT-V8 and the invasively measured LVED across all LBB pacing subtypes (RVSP, DSP, LVSP, ns/sLBBP) with a mean bias of 2.5ms; and (2) NDAT-V8 demonstrated superior performance as compared measurements that can be obtained from the currently available leads (V6-RWPT and NDAT-V6), for both intrinsic and all LBBP pacing subtypes. This correlation establishes a strong foundation for future studies evaluating NDAT-V8 as a potential tool to guide effective ventricular resynchronization during LBBAP.

Previous work on the NDAT-V8 has demonstrated its value as a non-invasive marker of delayed activation in the left ventricular posterolateral wall, showing an almost perfect correlation with the invasively measured QLV interval obtained from the LV lead in both LBBB and IVCD patients.(11) LV leads positioned outside the posterolateral region demonstrated shorter QLV intervals, whereas the NDAT-V8 accurately reflected the *true* delay in posterolateral wall activation that would have been obtained with an improved lead location. Moreover, NDAT-V8 was able to discriminate IVCD patients with significant posterolateral wall delay who were likely to derive substantial benefit from CRT, while also identifying Strauss-positive LBBB patients in whom such delay was absent.

Despite growing enthusiasm for LBBAP as a strategy for CRT in HF patients, a more individualized approach is warranted, as a “one-size-fits-all” strategy may not always be optimal.(10) The CSPOT study prospectively enrolled 48 HF patients who underwent implantation of both LBBAP and LV leads with simultaneous invasive hemodynamic assessment.(9) Despite expert implantation and precise lead positioning at the left subendocardial septum, deep septal pacing (DSP) was achieved in a surprisingly large proportion (44%) of cases—defined by the absence of a terminal R/r in V1 despite confirmed LV endocardial penetration and fulfillment of conventional activation criteria for LBB capture. This so-called *functional* DSP, attributed to severe conduction system slowing limiting effective activation, was associated with longer V6-RWPT, less QRS narrowing, and inferior hemodynamic performance compared to BiVP. Importantly, in patients with a baseline QRS duration >171 ms (in both LBBB and IVCD) or in whom only DSP was achieved, the addition of an LV lead to obtain LOT-CRT yielded significant further acute hemodynamic improvement. These findings suggest that a substantial subset of CRT candidates undergoing LBBAP derive additional benefit from implantation of an LV lead for LOT-CRT. Notably, more recent results demonstrated a significantly greater improvement in LVEF with LOT-CRT compared to a matched BiVP cohort (16.1% vs. 6.1%; *p* < 0.01).

However, determining when to implant an additional LV lead remains an ongoing clinical challenge in LBBAP. Several electrocardiographic and procedural characteristics observed during LBBAP implantation have been proposed to guide this decision.(19) The presence of either an IVCD (favoring additional LV lead or LV lead alone) versus isolated LBBB (which may favor CSP) has been used.(20) A larger baseline QRS duration suggests accompanying IVCD and likely need for an additional LV lead.(9) The presence of septal scar may also limit the ability to achieve a suitable response as a result of inability to advance the lead,(22) or presence of slow conduction when the LBB is reached (so-called functional DSP), for which an additional LV lead would be required.(9) Similarly, the inability to correct the LBBB with temporary His bundle pacing suggests presence of distal conduction disease that may not be amendable to LBBP. Other useful tools include the presence of persistently delayed LV lateral wall activation during coronary sinus mapping after LBBAP, non-invasive mapping using ECG imaging or ultra-high-frequency ECG, and lack of appropriate blood pressure response to LBBAP alone.(19)

Nevertheless, an individualized decision-making framework that quantifies benefit for a given patient remains lacking. This need for personalization is particularly relevant with the anticipated introduction of integrated shock leads within LBBAP systems, which will soon become commercially available.

The present study highlights the clinical relevance of the correlation of NDAT-V8 with LVED, also referred to as QLV. By providing a physiologically grounded marker of activation timing, NDAT-V8 may help to personalize resynchronization therapy. As the first study to systematically evaluate NDAT-V8 in relation to LVED across multiple pacing modalities during LBBAP, these findings represent an important step toward establishing individualized, physiology-based frameworks for resynchronization therapy.

### Limitations

A potential limitation of implementing V8 is the reproducibility of electrode placement and NDAT interpretation in less experienced centers or across different operators. Nonetheless, low inter-observer variability has been observed in previous studies(11), coupled with the ease with which nursing staff integrated V8 into routine workflow, supports its feasibility for broader adoption in clinical practice. Not all LBBAP subtypes were captured for all patients given either their absence (eg. transition from NS to S vs NS to LVSP) or omission. However despite this, there was such excellent consistency of correlation across all LBBAP subtypes.

### Conclusion

The NDAT-V8 provides a non-invasive surrogate for the LV posterolateral wall delay during LBBAP, demonstrating superior performance compared with NDAT-V6 and the V6-RWPT. Having established this correlation, incorporating lead V8 into clinical practice may assist in identifying patients who could benefit from the addition of an LV lead, thereby enabling more individualized and effective cardiac resynchronization therapy.

## Data Availability Statement

The data underlying this article will be shared on reasonable request to the corresponding author.

## Acknowledgments

JJ and VE are recipients of Clinical Research Scholar Awards from the Fonds de recherche du Québec-Santé (FRQS). UN is a recipient of Dutch Heart Foundation (Dekker 2021-T016, Dekker 2024–0559, Public-Private Partnership 2024–0477) and the Dutch Research Council (ZonMW Veni 2025).

---

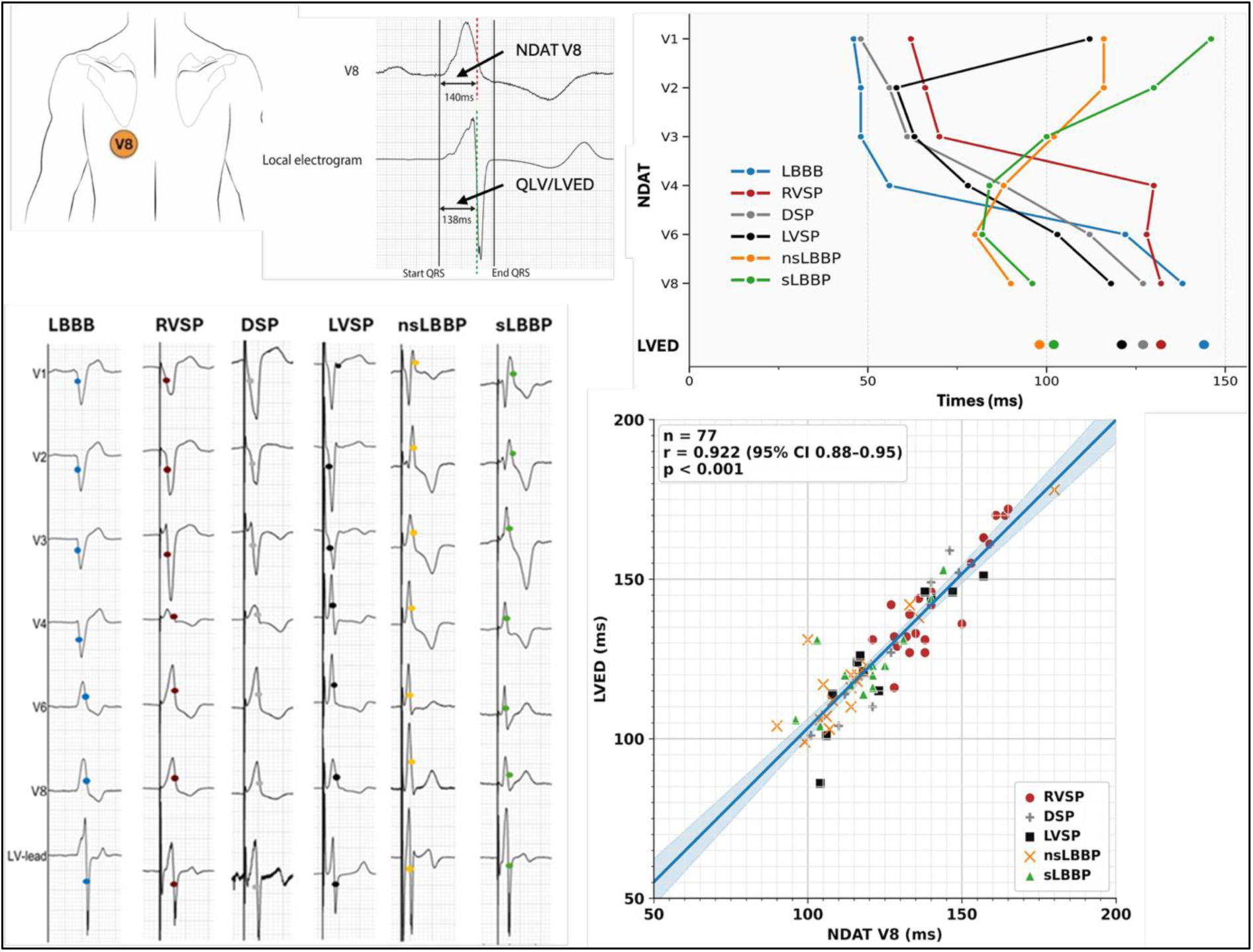
Graphical abstract: The negative derivative activation time (NDAT) of the surface electrode positioned at V8 closely corresponds to the left ventricular electrical delay to the left ventricular lead within the coronary sinus during intrinsic rhythm, and during all left bundle branch area pacing subtypes (right ventricular septal pacing (RVSP), deep septal pacing (DSP), left ventricular septal pacing (LVSP), non-selective left bundle branch pacing (nsLBBP), and selective left bundle branch pacing (sLBBP). Abbreviations: LBBB: left bundle branch block

